# Surgical Intervention for Paediatric Infusion-Related Extravasation Injury: A Systematic Review

**DOI:** 10.1101/19008664

**Authors:** MR Little, S Dupré, JCR Wormald, MD Gardiner, C Gale, A Jain

## Abstract

**Objectives:** This systematic review aims to assess the quality of literature supporting surgical interventions for paediatric extravasation injury and to determine and summarize their outcomes.

**Methods:** We performed a systematic review by searching Ovid MEDLINE and EMBASE as well as AMED, CINAHL, Cochrane Central Register of Controlled Trials (CENTRAL), Cochrane Database of Systematic Reviews and clinicaltrials.gov from inception to February 2019. All studies other than case reports were eligible for inclusion if the population was younger than 18 years old, there was a surgical intervention aimed at treating extravasation injury and they reported on outcomes. Risk of bias was graded according to the National Institutes of Health (NIH) study quality assessment tools.

**Results:** 26 studies involving 728 children were included – one before-and-after study and 25 case series. Extravasation injuries were mainly confined to skin and subcutaneous tissues but severe complications were also encountered, including amputation (one toe and one below elbow). Of the surgical treatments described, the technique of multiple puncture wounds and instillation of saline and/or hyaluronidase was the most commonly used. However, there were no studies in which its effectiveness was tested against another treatment or a control and details of functional and aesthetic outcomes were generally lacking.

**Conclusion:** There is a lack of high quality evidence to support treatment of extravasation injury in children. A definitive trial of extravasation injuries, or a centralized extravasation register using a universal grading scheme and core outcome set with adequate follow-up, are required to provide evidence to guide clinician decision-making.

**Strengths and Limitations:**

- A systematic review was performed according to PRISMA guidelines and registered on PROSPERO
- Two authors used a bespoke inclusion/exclusion form to independently assess study eligibility
- Studies were eligible for inclusion if the population was younger than 18 years old, if there was a surgical intervention aimed at treating extravasation injury in any setting and if they reported on short- or long-term outcomes
- Two researchers also independently assessed the included studies’ risk of methodological bias using the National Institutes of Health (NIH) study quality assessment tools
- 18 years old may represent a relatively arbitrary cut-off age to differentiate between ‘paediatric’ and ‘adult’ in terms of extravasation injury

## Introduction

Extravasation is the inadvertent leakage of a vesicant solution from its intended vascular pathway, commonly a peripheral or central vein, into the surrounding tissue with the potential to harm a patient. A vesicant is any medicine or fluid with the potential to cause blisters, severe tissue injury or necrosis if there is leakage. ^1^ If left untreated, extravasations may result in delayed healing, scarring and functional morbidity, including amputation in severe cases.

Children requiring intravenous (IV) therapy often have multiple risk factors for extravasation injuries, and neonates are at even greater risk of more serious injury due to poor venous integrity, capillary leakage, and expandable subcutaneous tissue that accommodates relatively large volumes of fluid. ^2^

The reported incidence of extravasation in adult and paediatric inpatients ranges between 0.1% - 6.5% across studies from the UK, USA and Canada, but inconsistent reporting and documentation means that the true figure is likely to be higher. ^2–4^

Multiple treatments have been described. The “Gault technique” ^5^ utilizes multiple small incisions made around the injury site. These are subsequently used to instill saline and/or hyaluronidase to wash out the extravasation fluid. This method remains a commonly described technique for the treatment of more severe paediatric extravasations. However, there is a lack of high quality comparative studies in which treatments are evaluated against each other and/or controls. It is therefore not known which of these treatments are effective, harmful or at the very least, necessary; a conclusion supported by a recent scoping review of interventions. ^6^ That review highlighted the fact that the lack of consensus extended to published guidelines. Their survey also showed wide variation in the frequency of use of the saline washout techniques in neonatal units across the National Health Service (NHS) in the UK. This systematic review aims to evaluate the quality of the literature and to summarize the published outcome data of the surgical management of paediatric extravasation injuries.

## Methods

We performed a systematic review of original clinical studies or systematic reviews (excluding case reports) of paediatric extravasation injury, reported in accordance with the Preferred Reporting Items for Systematic Reviews and Meta-Analyses (PRISMA) ^7^ statement and using methodology outlined in the Cochrane Handbook for Systematic Reviews of Interventions ^8^ where applicable. The protocol was developed prospectively, locally peer-reviewed and registered on the PROSPERO database (CRD42016045647).

### Search methods

Studies were identified through a systematic literature search in Ovid MEDLINE and Ovid EMBASE as well as AMED, CINAHL, Cochrane Central Register of Controlled Trials (CENTRAL), Cochrane Database of Systematic Reviews and clinicaltrials.gov from inception to February 2019. To identify grey literature, Web of Science was searched using bespoke searches. Internet-wide searches using Google were employed to identify clinical guidelines describing the management of extravasation in the UK. All of these strategies comprised modifications of a common set of terms to identify the relevant clinical area. Key search terms in this common set included the Medical Subject Headings (MeSH) “extravasation of diagnostic and therapeutic materials”, “infant”, “child”, and “preschool”. These MeSH terms were combined with appropriate Boolean operators [Supplementary 1]. The index search terms were then combined with free text terms to identify appropriate clinical studies. No language restrictions were imposed.

The search results were merged and, after screening, duplicate citations were discarded along with studies unrelated to the research objective.

### Criteria for selecting studies

Study selection criteria were determined in advance during the protocol stage. Two authors used a bespoke inclusion/exclusion form to independently assess eligibility of the title and abstracts of the search results. [Supplementary 2] Randomized Control Trials (RCTs), quasi-experimental and cross-sectional studies and case-series were eligible for inclusion if the population was younger than 18 years old, if there was a surgical intervention aimed at treating extravasation injury in any setting and if they reported on short- or long-term outcomes. Any disparities regarding inclusion of articles were discussed between the authors and a joint decision was made based on the inclusion criteria.

### Assessment of Bias

Two researchers also independently assessed the included studies’ risk of methodological bias using the National Institutes of Health (NIH) study quality assessment tools ^9^ for “case series” and for “before-after studies with no control group” – the two types of included studies. Discrepancies were resolved by discussions with the other authors.

## Results

### Search strategy

Our search strategy identified a total of 1,966 research articles of which 220 were potentially relevant to the research question. Of the 220 papers 185 did not meet the inclusion criteria. Thirty-five papers were deemed eligible for inclusion after studies in which conservative management or no intervention were excluded. The requisite data were immediately available in 26 study reports ^4,10,19–28,11,29–34,12–18^ and the authors of the remaining nine studies ^5,35–42^ were contacted twice with additional requests, mainly for exclusively paediatric extravasation data, and given at least two months to reply. Unfortunately, none of the authors were able to provide these data and those studies were excluded. The data were extracted from the remaining 26 studies [Supplementary PRISMA flow-diagram] using a pre-specified (review-specific) proforma [Supplementary 3] by two researchers, with discrepancies resolved by a third.

### Study Characteristics

The included studies were published between 1975 and 2018. The mean sample size was 32 with a range of 4-147 participants. One study was a before-and-after comparative study, ^31^ two were prospective case series, ^19,24^ and the remaining 23 were retrospective case series. The follow-up periods were not reported in 14 included studies. In the other studies, the follow-up periods ranged from three days to 15 months [Table 1].

**Table 1.** Included Studies.

| Rf | Author | Year | Country | Title | n | Methods | Period | Mean follow-up |
| --- | --- | --- | --- | --- | --- | --- | --- | --- |
| 4 | Murphy | 2017 | Australia | Extravasation injury in a paediatric population | 43 | Retrospective Series | - | - |
| 10 | Falcone | 1989 | USA | Nonoperative management of full-thickness intravenous extravasation injuries in premature neonates using enzymatic debridement | 15 | Retrospective Series | - | 6 months |
| 19 | Ghanem | 2015 | UK | Childhood extravasation injuries: improved outcome following the introduction of hospital-wide guidelines | 48 | Prospective Series | 1 year | - |
| 20 | Kostogloudis | 2015 | Greece | Severe Extravasation Injuries in Neonates: A Report of 34 Cases | 34 | Retrospective Series | 2 years | 15 months |
| 21 | Sung | 2016 | Korea | Nonoperative Management of Extravasation Injuries Associated With Neonatal Parenteral Nutrition Using Multiple Punctures and a Hydrocolloid Dressing. | 12 | Retrospective Series | 4 years | 10 months |
| 22 | Odom | 2018 | USA | Peripheral Infiltration and Extravasation Injury Methodology: A Retrospective Study. | 147 | Retrospective Series | - | - |
| 23 | Compañía | 2017 | Spain | Lesions Associated With Calcium Gluconate Extravasation | 4 | Retrospective Series | - | 2 months |
| 24 | Harris | 2001 | UK | Limiting the damage of iatrogenic extravasation injury in neonates | 56 | Prospective Series | 3 years | - |
| 25 | Andrés | 2006 | Spain | Treatment protocol for extravasation lesions | 15 | Retrospective Series | 6 years | - |
| 26 | Linder | 1983 | USA | Management of extensive doxorubicin hydrochloride extravasation injuries | 18 | Retrospective Series | - | 3 months |
| 27 |  | 1985 |  | Prevention of extravasation injuries secondary to doxorubicin |  |  |  |  |
| 28 | Upton | 1979 | USA | Major intravenous extravasation injuries | 7 | Retrospective Series | 10 years | - |
| 11 | Casanova | 2001 | France | Emergency treatment of accidental infusion leakage in the newborn: Report of 14 cases | 14 | Retrospective Series | - | - |
| 29 | von Heimburg | 1998 | Germany | Early and late treatment of iatrogenic injection damage | 5 | Retrospective Series | - | - |
| 30 | Weiss | 1975 | Israel | Localized necrosis of scalp in neonates due to calcium gluconate infusions: a cautionary note | 4 | Retrospective Series | - | - |
| 31 | Hanrahan | 2013 | USA | Hyaluronidase for treatment of intravenous extravasations: implementation of an evidence-based guideline in a pediatric population | 113 | Before-and-after study | 4 years | - |
| 32 | Boyar | 2018 | USA | Efficacy of Dehydrated Human Amniotic Membrane Allograft for the Treatment of Severe Extravasation Injuries in Preterm Neonates | 4 | Retrospective Series | - | 1-2 months |
| 33 | Yan | 2017 | China | Incidence, risk factors and treatment outcomes of drug extravasation in pediatric patients in China | 18 | Retrospective Series | 6 months | 1– 5 months |
| 34 | Myers | 2018 | USA | Managing intravenous infiltration injuries in the neonatal intensive care unit | 28 | Retrospective Series | 7 years | - |
| 12 | Cochran | 2002 | USA | Treatment of iodinated contrast material extravasation with hyaluronidase | 8 | Retrospective Series | 7 years | - |
| 13 | Cho | 2007 | Korea | Successful combined treatment with total parenteral nutrition fluid extravasation injuries in preterm infants | 5 | Retrospective Series | 4 months | - |
| 14 | Onesti | 2012 | Italy | The use of hyalomatrix PA in the treatment of extravasation affecting premature neonates | 26 | Retrospective Series | 6 years | 14 months |
| 15 | Firat | 2013 | Turkey | Management of extravasation injuries: a retrospective study | 13 | Retrospective Series | 2 years | - |
| 16 | Ching | 2014 | UK | Paediatric extravasation injuries: a review of 69 consecutive patients | 69 | Retrospective Series | 1 year | 3 days |
| 17 | Sivrioglu | 2014 | Turkey | Versajet hydrosurgery system in the debridement of skin necrosis after Ca gluconate extravasation: report of 9 infantile cases | 9 | Cohort Study | - | 1 year |
| 18 | Yan | 2014 | China | Treatment of cutaneous injuries of neonates induced by drug extravasation with hyaluronidase and hirudoid | 13 | Retrospective Series | 2 years | 3 months |

### Patient Characteristics

Across the 26 included studies, there were 728 children. The median age of the 68 children for whom the individual data was available was 0.66 months (interquartile range (IQR) = 5.05). The mean age of 14 months (range: neonatal to 17 years old) was calculated using the data for 322 children across 16 included studies. There was a small difference in sex, with 187 males (57%) and 142 females (43%) (data on sex was lacking for 399 children). 19 of the 26 included studies recorded the sites of the implicated cannula for 553 children. The most common site was the upper limb (n=343; 62%), followed by the lower limb (n=177; 32%) and then the scalp (n=21; 4%). Ten studies specifically stated whether the cannula was a peripheral (n=314; 98%) or central line (n=7; 2%). [Table 2]

**Table 2.**
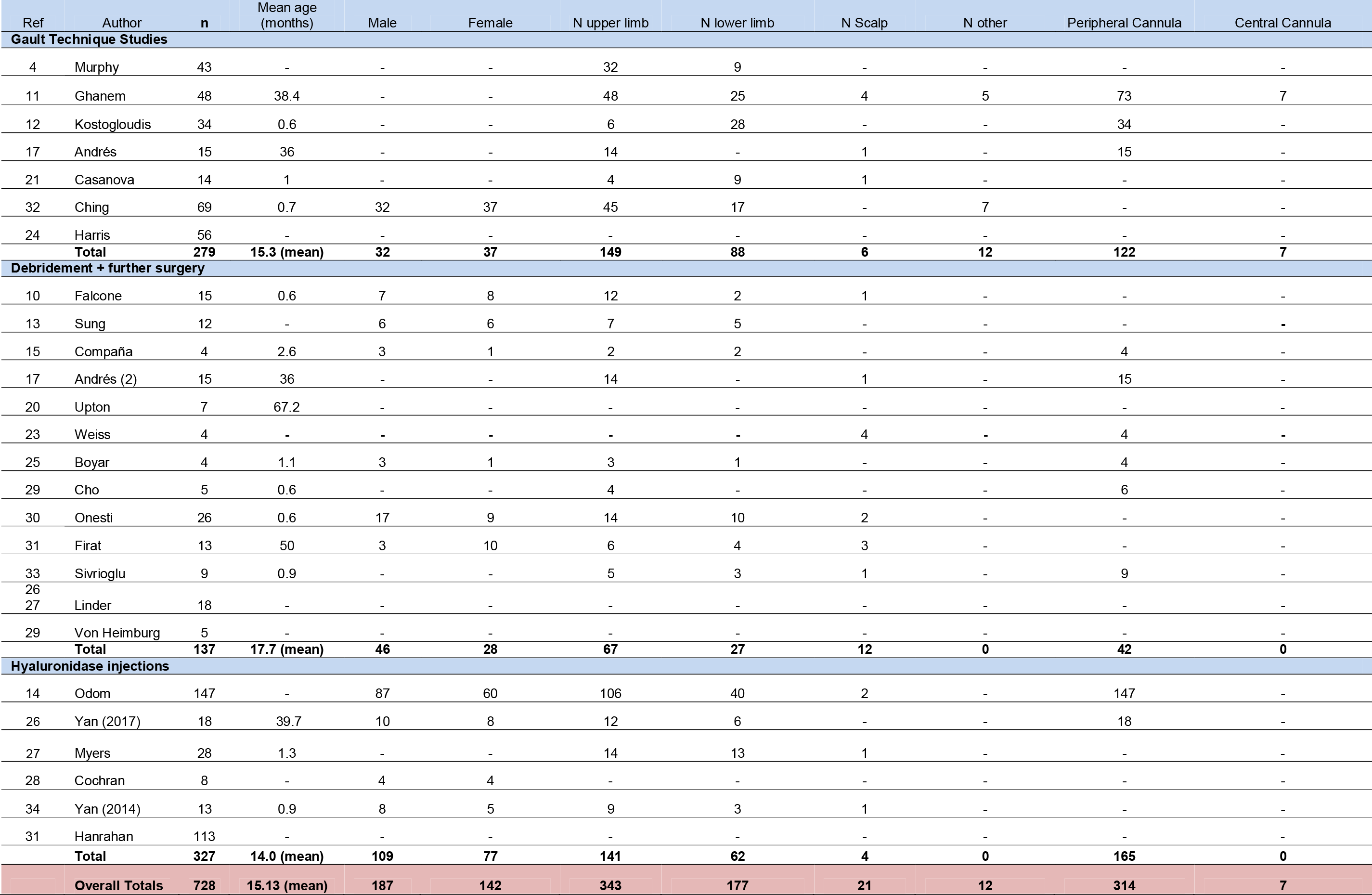
Demographics.

| Ref | Author | n | Mean age (months) | Male | Female | N upper limb | N lower limb | N Scalp | N other | Peripheral Cannula | Central Cannula |
| --- | --- | --- | --- | --- | --- | --- | --- | --- | --- | --- | --- |
| **Gault Technique Studies** |  |  |  |  |  |  |  |  |  |  |  |
| 4 | Murphy | 43 | - | - | - | 32 | 9 | - | - | - | - |
| 11 | Ghanem | 48 | 38.4 | - | - | 48 | 25 | 4 | 5 | 73 | 7 |
| 12 | Kostogloudis | 34 | 0.6 | - | - | 6 | 28 | - | - | 34 | - |
| 17 | Andrés | 15 | 36 | - | - | 14 | - | 1 | - | 15 | - |
| 21 | Casanova | 14 | 1 | - | - | 4 | 9 | 1 | - | - | - |
| 32 | Ching | 69 | 0.7 | 32 | 37 | 45 | 17 | - | 7 | - | - |
| 24 | Harris | 56 | - | - | - | - | - | - | - | - | - |
|  | **Total** | **279** | **15.3 (mean)** | **32** | **37** | **149** | **88** | **6** | **12** | **122** | **7** |
| **Debridement + further surgery** |  |  |  |  |  |  |  |  |  |  |  |
| 10 | Falcone | 15 | 0.6 | 7 | 8 | 12 | 2 | 1 | - | - | - |
| 13 | Sung | 12 | - | 6 | 6 | 7 | 5 | - | - | - | - |
| 15 | Compañía | 4 | 2.6 | 3 | 1 | 2 | 2 | - | - | 4 | - |
| 17 | Andrés (2) | 15 | 36 | - | - | 14 | - | 1 | - | 15 | - |
| 20 | Upton | 7 | 67.2 | - | - | - | - | - | - | - | - |
| 23 | Weiss | 4 | - | - | - | - | - | 4 | - | 4 | - |
| 25 | Boyar | 4 | 1.1 | 3 | 1 | 3 | 1 | - | - | 4 | - |
| 29 | Cho | 5 | 0.6 | - | - | 4 | - | - | - | 6 | - |
| 30 | Onesti | 26 | 0.6 | 17 | 9 | 14 | 10 | 2 | - | - | - |
| 31 | Firat | 13 | 50 | 3 | 10 | 6 | 4 | 3 | - | - | - |
| 33 | Sivrioglu | 9 | 0.9 | - | - | 5 | 3 | 1 | - | 9 | - |
| 26 | Linder | 18 | - | - | - | - | - | - | - | - | - |
| 27 |  |  |  |  |  |  |  |  |  |  |  |
| 29 | Von Heimburg | 5 | - | - | - | - | - | - | - | - | - |
|  | **Total** | **137** | **17.7 (mean)** | **46** | **28** | **67** | **27** | **12** | **0** | **42** | **0** |
| **Hyaluronidase injections** |  |  |  |  |  |  |  |  |  |  |  |
| 14 | Odom | 147 | - | 87 | 60 | 106 | 40 | 2 | - | 147 | - |
| 26 | Yan (2017) | 18 | 39.7 | 10 | 8 | 12 | 6 | - | - | 18 | - |
| 27 | Myers | 28 | 1.3 | - | - | 14 | 13 | 1 | - | - | - |
| 28 | Cochran | 8 | - | 4 | 4 | - | - | - | - | - | - |
| 34 | Yan (2014) | 13 | 0.9 | 8 | 5 | 9 | 3 | 1 | - | - | - |
| 31 | Hanrahan | 113 | - | - | - | - | - | - | - | - | - |
|  | **Total** | **327** | **14.0 (mean)** | **109** | **77** | **141** | **62** | **4** | **0** | **165** | **0** |
|  | **Overall Totals** | **728** | **15.13 (mean)** | **187** | **142** | **343** | **177** | **21** | **12** | **314** | **7** |

Of the 16 studies that provided data on comorbidities for 167 children, prematurity was the most common comorbidity in 11 (n=121; 72%). Malignancy (n=25; 15%) was the main comorbidity in three studies.

**Table 3.**
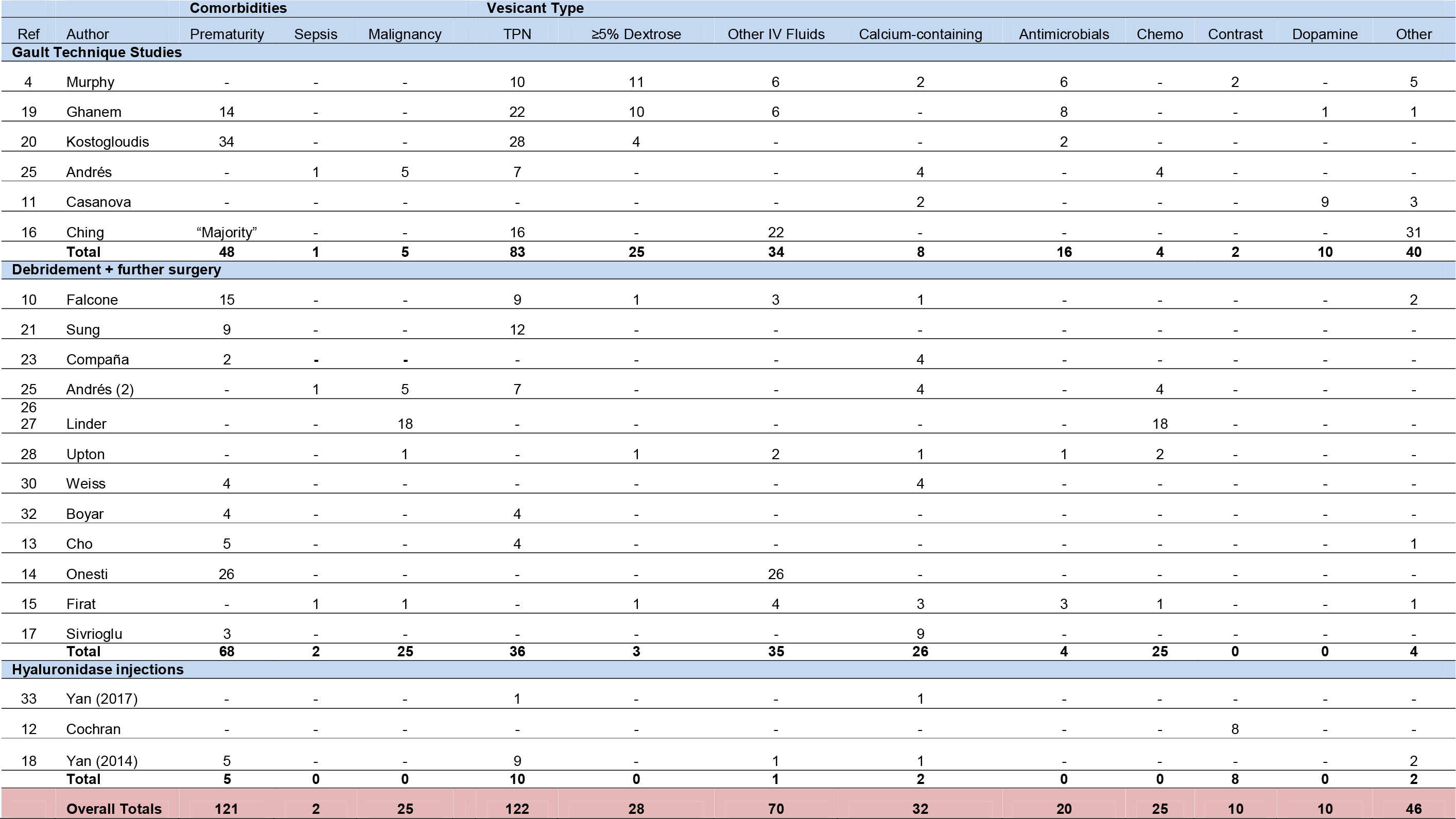
Comorbidities and extravasated materials in those papers that recorded them.

Of the 20 studies that reported on them, the most commonly extravasated substances affecting 363 children were [Table 3]:

1. Total Parenteral Nutrition (TPN) – 122 children (34%); most common in eight studies.
2. Intravenous maintenance fluids other than those specifically listed – 70 children (19%); most common in three studies. Unfortunately the data was not available for the outcomes of different types of maintenance fluids so they have been grouped together.
3. Calcium-containing products – 32 children (9%); most common in four studies.
4. Dextrose of concentration ≥5% - 28 children (8%); most common in one study.
5. Intravenous antimicrobials – 20 children (6%); most common in no studies. Of those 20 extravasations, five were due to Flucloxacillin with one injury taking 60 days to heal and five were due to unspecified cephalosporin antibiotics, three of which required skin grafting. One case of severe contractures and extensor loss on the dorsum of a child’s hand was due to an unspecified tetracycline antibiotic. There were no individual outcomes recorded for the seven extravasations of Aciclovir, the single extravasations of Gentamicin, Ganciclovir or the remaining extravasations involving Flucloxacillin and cephalosporin antibiotics.

### Interventions

Uncertainty remains over whether there is a difference in the outcomes of children who are treated conservatively or surgically and the case series that exist cover a number of treatments for a range of injuries from swelling to full-thickness defects. However, a recent scoping review found that the outcomes used and results detailed were generally limited, ^6^ making it difficult for clinicians to rely upon those studies in deciding who to treat conservatively. This is why we have focused solely on surgical interventions.

#### The Gault Method

279 children with an average age of 15.3 months were involved in the seven studies looking at the Gault method. No patients had skin necrosis at presentation.

107 were treated using the Gault method with saline, of which nine (8%) developed full thickness skin injury / skin necrosis and one (1%) developed compartment syndrome requiring fasciotomies. Of those with necrosis, six required no surgical intervention and three were debrided and treated with artificial dermis. [Table 4]

**Table 4.**
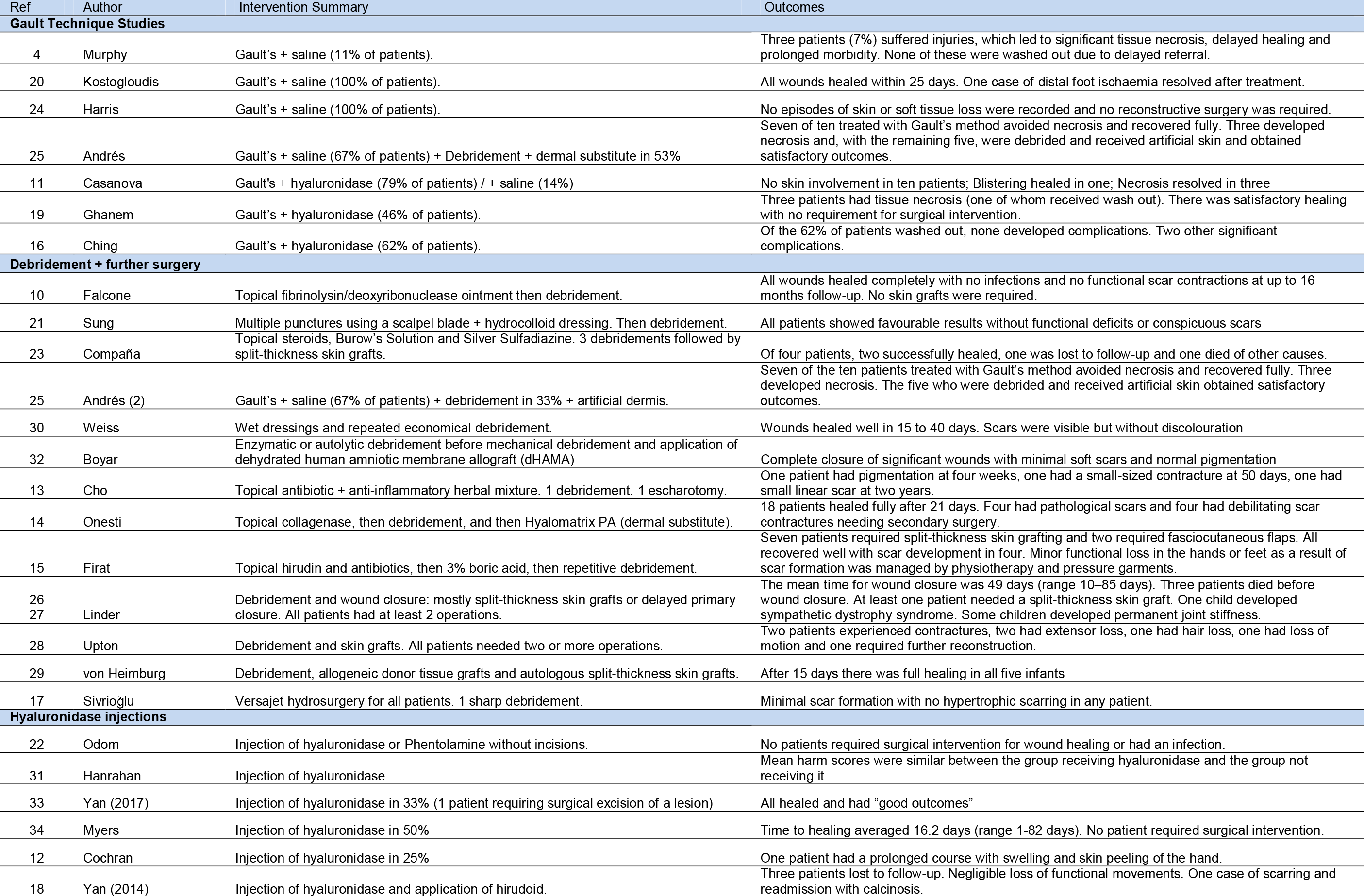
Interventions.

**Table 5.**
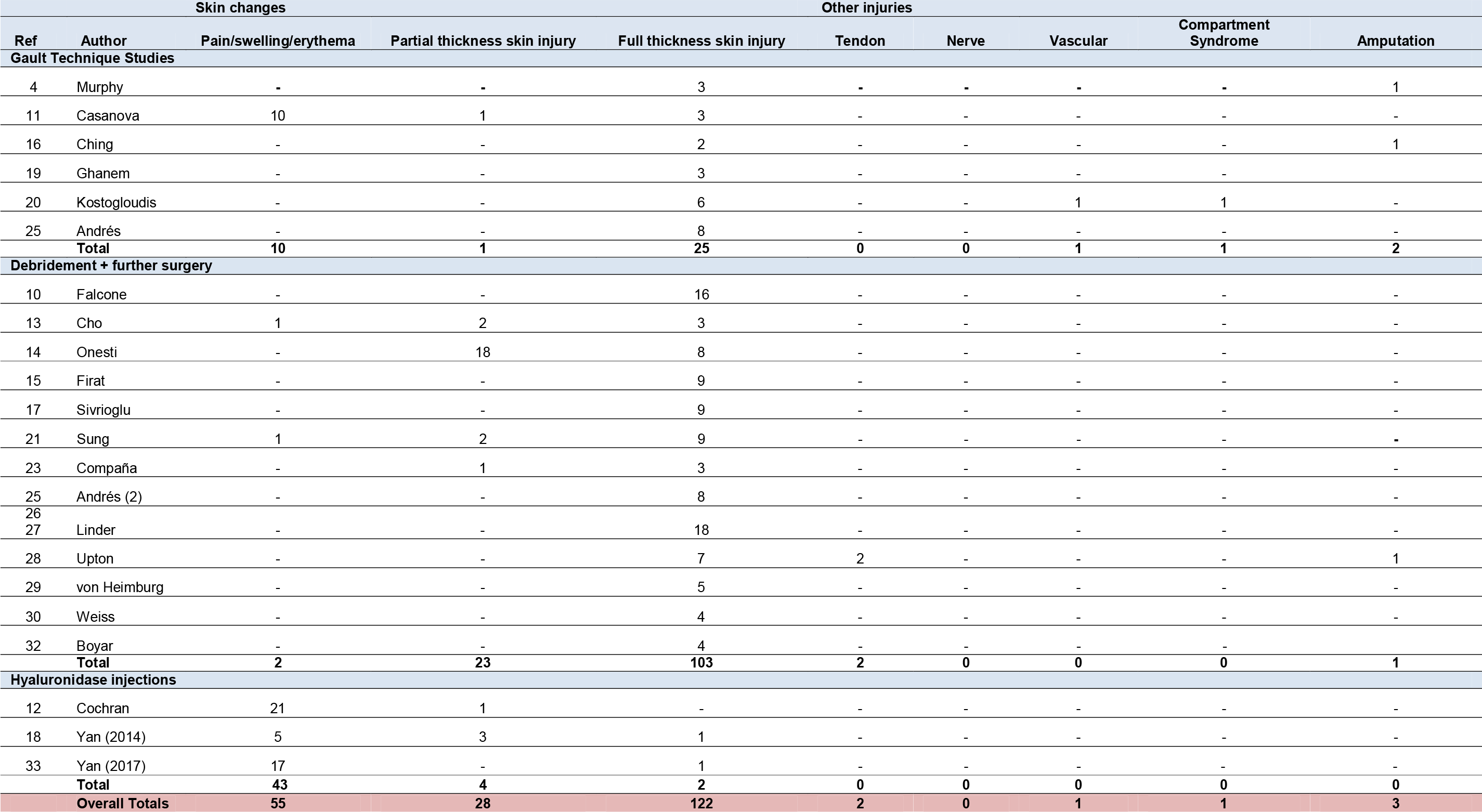
Injuries in those papers that recorded them.

**Table 6.**
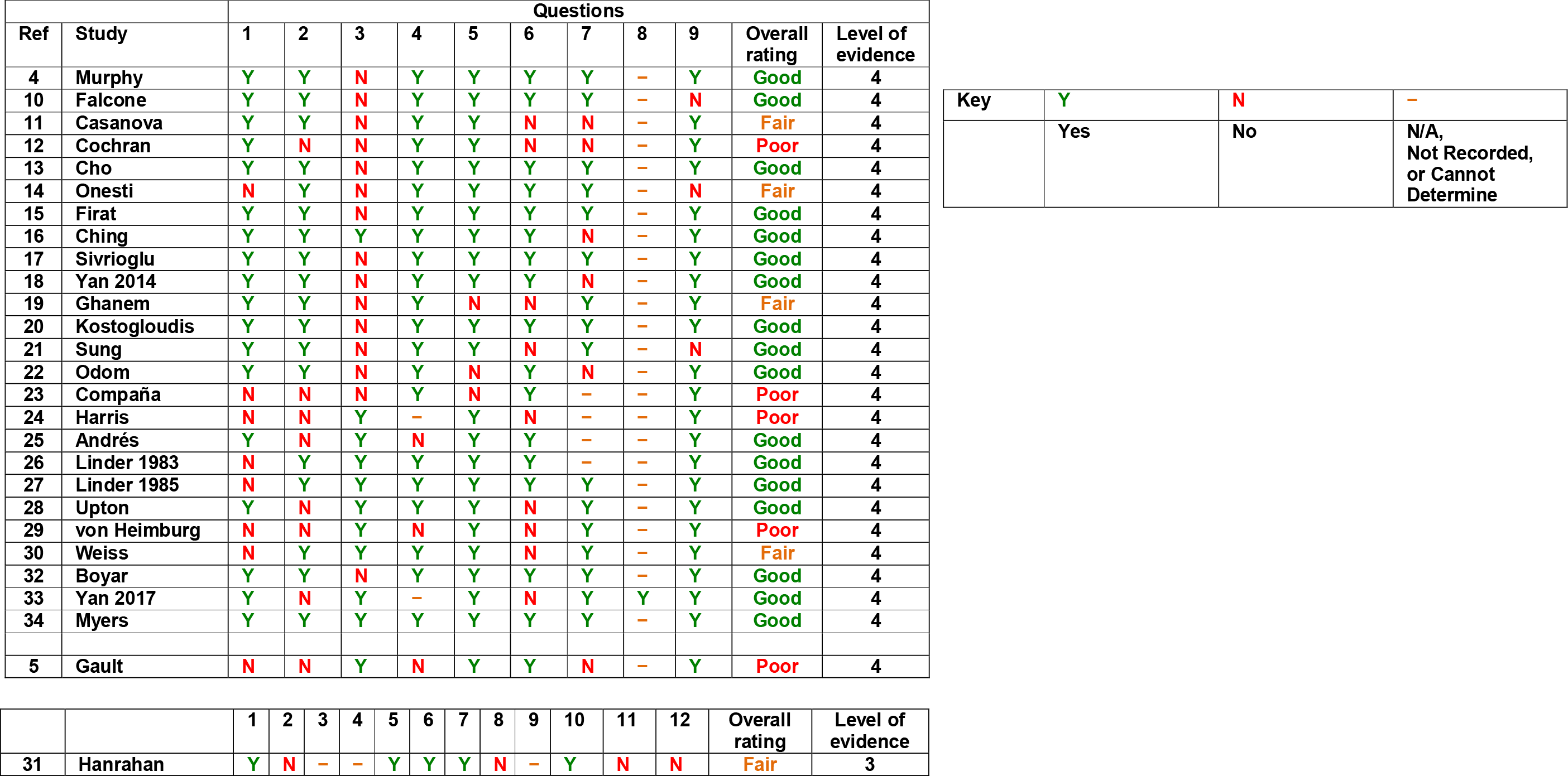
Risk of Bias for Included Studies.

| Ref | Study | Questions |  |  |  |  |  |  |  |  | Overall rating | Level of evidence |
| --- | --- | --- | --- | --- | --- | --- | --- | --- | --- | --- | --- | --- |
|  |  | 1 | 2 | 3 | 4 | 5 | 6 | 7 | 8 | 9 |  |  |
| 4 | Murphy | Y | Y | N | Y | Y | Y | Y | – | Y | Good | 4 |
| 10 | Falcone | Y | Y | N | Y | Y | Y | Y | – | N | Good | 4 |
| 11 | Casanova | Y | Y | N | Y | Y | N | N | – | Y | Fair | 4 |
| 12 | Cochran | Y | N | N | Y | Y | N | N | – | Y | Poor | 4 |
| 13 | Cho | Y | Y | N | Y | Y | Y | Y | – | Y | Good | 4 |
| 14 | Onesti | N | Y | N | Y | Y | Y | Y | – | N | Fair | 4 |
| 15 | Firat | Y | Y | N | Y | Y | Y | Y | – | Y | Good | 4 |
| 16 | Ching | Y | Y | Y | Y | Y | Y | N | – | Y | Good | 4 |
| 17 | Sivrioglu | Y | Y | N | Y | Y | Y | Y | – | Y | Good | 4 |
| 18 | Yan 2014 | Y | Y | N | Y | Y | Y | N | – | Y | Good | 4 |
| 19 | Ghanem | Y | Y | N | Y | N | N | Y | – | Y | Fair | 4 |
| 20 | Kostogloudis | Y | Y | N | Y | Y | Y | Y | – | Y | Good | 4 |
| 21 | Sung | Y | Y | N | Y | Y | N | Y | – | N | Good | 4 |
| 22 | Odom | Y | Y | N | Y | N | Y | N | – | Y | Good | 4 |
| 23 | Compañá | N | N | N | Y | N | Y | – | – | Y | Poor | 4 |
| 24 | Harris | N | N | Y | – | Y | N | – | – | Y | Poor | 4 |
| 25 | Andrés | Y | N | Y | N | Y | Y | – | – | Y | Good | 4 |
| 26 | Linder 1983 | N | Y | Y | Y | Y | Y | – | – | Y | Good | 4 |
| 27 | Linder 1985 | N | Y | Y | Y | Y | Y | Y | – | Y | Good | 4 |
| 28 | Upton | Y | N | Y | Y | Y | N | Y | – | Y | Good | 4 |
| 29 | von Heimburg | N | N | Y | N | Y | N | Y | – | Y | Poor | 4 |
| 30 | Weiss | N | Y | Y | Y | Y | N | Y | – | Y | Fair | 4 |
| 32 | Boyar | Y | Y | N | Y | Y | Y | Y | – | Y | Good | 4 |
| 33 | Yan 2017 | Y | N | Y | – | Y | N | Y | Y | Y | Good | 4 |
| 34 | Myers | Y | Y | Y | Y | Y | Y | Y | – | Y | Good | 4 |
| 5 | Gault | N | N | Y | N | Y | Y | N | – | Y | Poor | 4 |

|  |  |  |  |
| --- | --- | --- | --- |
| Key | Y | N | – |
|  | Yes | No | N/A,<br>Not Recorded,<br>or Cannot<br>Determine |

|  |  | 1 | 2 | 3 | 4 | 5 | 6 | 7 | 8 | 9 | 10 | 11 | 12 | Overall rating | Level of evidence |
| --- | --- | --- | --- | --- | --- | --- | --- | --- | --- | --- | --- | --- | --- | --- | --- |
| 31 | Hanrahan | Y | N | – | – | Y | Y | Y | N | – | Y | N | N | Fair | 3 |

76 were treated using the Gault method with hyaluronidase, of which three (4%) developed skin necrosis and were all managed conservatively.

One child received only hyaluronidase injections and developed necrosis, which healed spontaneously.

95 children received no treatment, of which ten (11%) developed necrosis (one of whom went on to lose three toes), three (3%) developed an associated infection, one (1%) had an ischaemic toe requiring amputation and one (1%) developed calcinosis cutis.

#### Debridement and further surgery

There were 137 children with an average age of 17.7 months across the 14 studies in the debridement and further surgery group.

Nine studies (98 children) used management plans comprising debridement and further surgery in combination with conservative measures, details of which are shown in Table 4. ^10,13–15,21,23,25,30,32^ Of those 98 children, 34 (35%) had established full thickness skin injury / skin necrosis at presentation and 28 (29%) developed necrosis after conservative measures. 84 children (86%) underwent mechanical debridement as part of their treatment. The vast majority of children’s wounds healed with minimal scarring and no functional deficit but four (4%) had debilitating scar contractures necessitating further surgery, ten (10%) required skin grafts, and two (2%) required fasciocutaneous flaps.

The other five studies (39 children) used only debridement and further surgery. All of those 39 children had full thickness skin injury / skin necrosis at presentation. 30 (77%) underwent mechanical debridement, of whom 22 (56%) required further operations including skin grafts and fasciocutaneous flaps and two developed contractures. The remaining nine children (23%), all from one study, underwent Versajet hydrosurgery with good healing in all children and one requiring sharp debridement.

In total, of the 137 children in the debridement and further surgery group, 103 (75%) developed skin necrosis, all of whom (in addition to some of those who were yet to develop necrosis, total n=123 (89%)) received either mechanical or water jet debridement. At least 38 children (28%) went on to require further operations.

#### Hyaluronidase injections

The final group of studies used hyaluronidase injections at the extravasation site without the need for incisions or washouts. This group comprised six studies and 327 children.^12,18,22,31,33,34^

97 children received hyaluronidase injections, one (1%) of whom had skin necrosis at presentation that resolved, and one (1%) developed necrosis requiring further surgery. Of the remaining 230 children that did not receive hyaluronidase, none developed skin necrosis at any stage, although reporting on the extent of skin damage and eventual outcomes across the studies was generally poor.

### Comparison of the interventions

#### The Gault Method

Among the studies included in this review, Ghanem et al ^19^ provides the closest approximation of a comparative study of the Gault method in children: 48 extravasations were diagnosed and children were divided into early (<24 hours, n=45) and later (>24 hours, n=3) referrals. Among early referrals, 22 were deemed to be ‘at risk’ injuries and received washout using Gault’s method. Skin necrosis occurred in one of 45 early referrals but in two of three later referrals. The conclusion of this study, that the incidence of necrosis is higher in the later referral group, is limited by low methodological quality including the use of an arbitrary 24-hour cut-off, the small size of the later referral group and a lack of data describing the types of vesicants. It is important to note that the necrosis resolved with conservative management in all three cases.

Of the 183 children treated with a variation of the Gault technique across the included studies, 12 (7%) developed skin necrosis (none had skin necrosis at presentation). This compares favourably to the 11% (10/95) of children who received no treatment across those same studies. However, 73% (69/95) of the untreated group were late referrals and may be expected to have worse outcomes related to time of presentation. The only study to apply the Gault technique in which plastic surgery review time and patient ages were similar was Ching, Wong and Milroy. ^16^ In that study, none of the 43 children treated with Gault’s technique and hyaluronidase developed any complications whereas there were three cases of associated infection, one ischaemic toe requiring amputation and one case of calcinosis cutis amongst the 26 children that received no treatment.

The degree of injury was only recorded for 36 (13%) of the 279 children in the included studies describing the Gault technique, of which 22 (8%) were full-thickness skin injuries.

#### Debridement and further surgery

73 children (53%) of the 137 in the debridement and further surgery studies had at least skin necrosis / full thickness skin injury at presentation In contrast to the Gault as well as the hyaluronidase injection studies, the degree of injury was documented for 93% of the children (128/137).

#### Hyaluronidase injections

One (0.3%) of the 327 children reported in studies describing hyaluronidase injections had skin necrosis at presentation, and one (0.3%) went on to develop it. The degree of injury was recorded for only 49 (15%) of the 327 children. The reporting of patient outcomes was particularly scarce in this group.

### Injuries

The types of injury were recorded for 212 of the 728 children across the included studies. Injuries were limited to swelling, pain or erythema (n=63; 30%), partial-thickness skin injury such as superficial blistering in others (n=29; 14%) and full-thickness skin injury such as necrosis in the largest number of children (n=110; 52%). However, these figures are skewed by the fact that 10 studies only looked at extravasation injuries causing at least skin necrosis. ^10,15,17,20,26–30,32^ One study described a transient episode of foot ischaemia which resolved with conservative management and one compartment syndrome of the left forearm requiring fasciotomies. ^20^ Two studies described toe amputations secondary to ischaemia. ^4,16^ One study of major injuries with full-thickness tissue loss had several casualties including a below elbow amputation and two episodes of extensor loss. ^28^ Table 5 demonstrates the huge range of injury severity encountered and the number of each in the different intervention groups.

### Millam’s grading system for extravasation injuries

Several of the included studies ^11,13,17,20,22,33^ make reference to the four stages of extravasation/infiltration severity, which was first published by Millam in 1988. ^43^ This focuses on presence of pain, erythema, oedema, capillary refill, skin temperature and breakdown and it has been proposed that stages III & IV may require intervention. ^44^

In one case series, ^33^ treatment was determined by the severity of extravasation injury based on a grading system similar to Millam’s. Of the 18 children, the seven with Grade I and II injuries were managed conservatively. Ten children had grade III injuries – five were managed conservatively but the other five who had also developed skin erythema received active treatment with hyaluronidase injections. The one child with a grade IV injury required excision of the lesion – the sole surgical intervention in that study – after having been injected with hyaluronidase. All wounds healed with ‘good outcomes’.

### Risk of Bias

The one before-and-after study ^31^ was evaluated using a separate tool to the other studies (which were all case series). It was given an overall ‘fair’ rating by both authors. The vast majority of the case series were rated by both authors as ‘good’ or ‘fair’ [Table 6]. All four of the studies rated as ‘poor’ lacked a clearly defined study population. Compaña, Míguez & de Toro Santos ^23^, Harris, Bradley & Moss ^24^, and von Heimburg & Pallua ^29^ did not clearly state their objective whilst Cochran, Bomyea & Kahn ^12^, Harris, Bradley & Moss and von Heimburg & Pallua lacked clearly-defined, valid and reliable outcome measures.Compaña, Míguez & de Toro Santos did not clearly describe the study’s intervention and von Heimburg & Pallua’s subject were not comparable. In combination, these factors led to the overall ‘poor’ rating of these four studies, meaning that their results were at high risk of methodological bias.

## Discussion

This paper describes a systematic review of characteristics and operative management of paediatric extravasation injuries. Our review highlighted three key issues. Firstly, surgical management is commonly reported in the literature in cases where there is significant soft tissue injury but as there are no comparative studies, it is unclear whether this is optimal. Secondly, there are no data to support one surgical management approach over another, or over no treatment. Thirdly, there are no data on the adverse events caused by the different surgical interventions.

A variation of the Gault method was used in the largest number of children to receive surgical treatment across the included studies. Gault ^4^ described a retrospective case series of 96 patients, ranging in age from newborn to 70 years, who were split into two groups: those seen within 24 hours of the extravasation and those seen later than this. Patients seen within 24 hours had treatment as per the Gault method while those seen after 24 hours had no treatment. The original Gault paper was not included in our review because there were no separate paediatric data available but a separate risk of bias assessment by two authors rated the study as ‘poor’ due to the lack of a clearly defined objective or population, subjects that were not comparable and an inadequate length of follow-up. [Table 6] Despite the inherent limitations in the original Gault study, the method he described is widely used, as demonstrated by this review where it was a commonly described technique to manage extravasation injuries. This approach is not without downsides: it is an invasive procedure with associated morbidity such as pain and possible tendon and nerve damage, and takes considerable time to perform.

### Evidence for the continuing use of the “Gault technique”

The degree of injury was recorded for 13% of the children included in the Gault technique studies, 93% of those in the debridement and further surgery group and 15% of those in the hyaluronidase injection group. These figures demonstrate some of the difficulties faced in drawing comparisons between different interventions, both due to the poor recording of injuries and because of the fact that accurate documentation of injury severity appears to be more consistently applied following more severe injuries.

It was not possible to perform pooled analysis from included studies of Gault and alternative techniques due to study heterogeneity and incomplete outcome reporting.

All but one of the included studies were case series and the one before-and-after study ^31^ primarily examined the effect of new extravasation treatment guidelines on knowledge of hyaluronidase, extravasation incident reporting and time from extravasation discovery to treatment. Therefore, even though they found similar harm scores for the hyaluronidase and the non-hyaluronidase groups, the results could not be relied upon to assess the effectiveness of the intervention.

In addition, inclusion criteria varied considerably, details of the injuries were poorly described, and no studies made direct comparison to another intervention. Finally, detailed outcome measures following interventions were generally lacking. In summarizing the included studies, it can be noted that despite widespread use, there is no high-quality evidence to support the Gault approach or its modifications.

### The future of extravasation management

A key conclusion from this review is that resolution of extravasation injuries is common regardless of the treatment applied. In the absence of data to support one treatment modality over another, it is imperative that high quality methodology randomized comparisons are undertaken; however, given the high rate of resolution with conservative treatment, any such trial will require a large, collaborative and multidisciplinary approach. Although many of the in the included studies required minimal intervention, full-thickness skin injury was commonplace; this has the potential for lifelong scarring in patients with an immature skin barrier such as children and preterm infants in particular. More serious complications were rare in but given that only a minority of studies reported on longer-term outcomes, these may underestimate the true burden of extravasation injuries. We suggest the creation of a core outcome set for extravasation injury treatment and that future studies use longer-term follow-up to monitor those outcomes.

The use of a universal grading scheme (such as Millam’s ^43^) to assess the initial extent of injuries would also aid the comparison between different interventions. However, this requires meticulous documentation of the appearance of the extravasated site as well as the extent of tissue damage.

This review was limited by the low methodological quality of included studies and inconsistent outcome reporting. Variability in baseline data, intervention data and outcome data precluded formal meta-analysis. These limitations prevent the authors from providing definitive clinical recommendations either for or against paediatric extravasation, based on the current evidence.

## Conclusion

There is a paucity of evidence to inform surgical treatment of paediatric and neonatal extravasation injuries. Gault’s technique is a commonly described approach, but we did not identify any evaluation of its effectiveness compared to other approaches or conservative management. A centralized extravasation register, use of a standardized and agreed grading scheme, development of a core outcome set and adequate follow-up of extravasation injuries are required to provide incidence and outcome data for this condition, and to inform much-needed, definitive trials of therapeutic approaches to extravasation injuries in children and neonates.

## Data Availability

There is no supplementary data online. The protocol was registered on the PROSPERO database (CRD42016045647) - link below.

## References

1. Doellman D, Hadaway L, Bowe-Geddes LA, et al. Infiltration and extravasation: Update on prevention and management. J Infus Nurs. 2009;32(4):203–211. doi:10.1097/NAN.0b013e3181aac042

2. Paquette V, McGloin R, Northway T, et al. Describing intravenous extravasation in children (DIVE study). Can J Hosp Pharm. 2011;64(5):340–345.

3. Wilkins C, Emmerson A. Extravasation injuries on regional neonatal units. Arch Dis Child Fetal neonatal Ed. 2004;89(3).

4. Murphy A, Gilmour R, Coombs C. Extravasation injury in a paediatric population. ANZ J Surg. 2017.

5. Gault D. Extravasation injuries. Br J Plast Surg. 1993;46(2):91.

6. Corbett M, Marshall D, Harden M, et al. Treatment of extravasation injuries in infants and young children: a scoping review and survey. Southampton (UK): NIHR Journals Library. Health Technol Assess (Rockv). 2018;22(46). doi:10.3310/hta22460

7. Moher D, Liberati A, Tetziaff J, et al. Preferred Reporting Items for Systematic Reviews and Meta-Analyses. The PRISMA Statement. PLoS Med. 2009;6(7):e1000097. doi:10.1371/journal.pmed1000097

8. Higgins J, Green S. Cochrane Handnook for Systematic Reviews of Interventions Version 5.1.0 [updated March 2011]. Cochrane Collab. 2011. http://handbook.cochrane.org.

9. NIH Study Quality Assessment Tools. https://www.nhlbi.nih.gov/health-topics/study-quality-assessment-tools. Accessed March 25, 2019.

10. Falcone P, Barrall D, Jeyarajah D, et al. Nonoperative management of full-thickness intravenous extravasation injuries in premature neonates using enzymatic debridementtle. Ann Plast Surg. 1989;22(2):146.

11. Casanova D, Bardot J, Magalon G. Emergency treatment of accidental infusion leakage in the newborn: Report of 14 cases. Br J Plast Surg. 2001;54(5):396–399.

12. Cochran T, Bomyea K, Kahn M. Treatment of iodinated contrast material extravasation with hyaluronidase. Acad Radiol. 2002;9.

13. Cho K, Lee S, Burm J, et al. Successful combined treatment with total parenteral nutrition fluid extravasation injuries in preterm infants. J Korean Med Sci. 2007;22(3):588.

14. Onesti M, Carella S, Maruccia M, et al. The use of hyalomatrix PA in the treatment of extravasation affecting premature neonates. Plast Reconstr Surg. 2012;129(1):219e–21e.

15. Firat C, Erbatur S, Aytekin A. Management of extravasation injuries: a retrospective study. J Plast Surg Hand Surg. 2013;47(1):60.

16. Ching L, Wong K, Milroy C. Paediatric extravasation injuries: a review of 69 consecutive patients. Int J Surg. 2014;12(10):1036.

17. Sivrioğlu N, Irkören S. Versajet hydrosurgery system in the debridement of skin necrosis after calcium gluconate extravasation: report of 9 infantile cases. Acta Orthop Traumatol Turc. 2014;48(1):6.

18. Yan Y, Fan Q, Li A, et al. Treatment of cutaneous injuries of neonates induced by drug extravasation with hyaluronidase and hirudoid. Iran J Pediatr. 2014;24(4):352–358.

19. Ghanem M, Mansour A, Exton R, et al. Childhood extravasation injuries: improved outcome following the introduction of hospital-wide guidelines. J Plast Reconstr Aesthetic Surg. 2015;68(4):505.

20. Kostogloudis N, Demiri E, Tsimponis A, et al. Severe Extravasation Injuries in Neonates: A Report of 34 Cases. Pediatr Dermatol. 2015;32(6):830.

21. Sung K-Y, Lee S-Y. Nonoperative Management of Extravasation Injuries Associated With Neonatal Parenteral Nutrition Using Multiple Punctures and a Hydrocolloid Dressing. Wounds A Compend Clin Res Pract. 2016;28(5):145–152.

22. Odom B, Lowe L, Yates C. Peripheral Infiltration and Extravasation Injury Methodology: A Retrospective Study. J Infus Nurs. 2018;41(4):247–252.

23. Pacheco Compaña F, Midón Míguez J, de Toro Santos F. Lesions Associated With Calcium Gluconate Extravasation. Ann Plast Surg. 2017;79(5):444–449.

24. Harris P, Bradley S, Moss A. Limiting the damage of iatrogenic extravasation injury in neonates. Plast Reconstr Surg. 2001;107(3):893–894. doi: https://doi.org/10.1097/00006534-200103000-00053

25. Andrés A, Burgos L, López Gutiérrez J, et al. Treatment protocol for extravasation lesions. Cir Pediatr. 2006;19(3):136–139.

26. Linder R, Upton J, Osteen R. Management of extensive doxorubicin hydrochloride extravasation injuries. J Hand Surg Am. 1983;8(1):32–38. doi: https://doi.org/10.1016/S0363-5023(83)80048-3

27. Linder R, Upton J. Prevention of extravasation injuries secondary to doxorubicin. Postgrad Med J. 1985;77(4):105-9,112,114. doi: https://doi.org/10.1080/00325481.1985.11698920

28. Upton J, Mulliken J, Murray J. Major intravenous extravasation injuries. Am J Surg. 1979;137(4):497–506. doi: https://doi.org/10.1016/0002-9610(79)90121-1

29. von Heimburg D, Pallua N. Early and late treatment of iatrogenic injection damage. Chirurg. 1998;69(12):1378–1382. doi: https://doi.org/10.1007/s001040050588

30. Weiss Y, Ackerman C, Shmilovitz L. Localized necrosis of scalp in neonates due to calcium gluconate infusions: a cautionary note. Pediatrics. 1975;56(6):1084–1086.

31. Hanrahan K. Hyaluronidase for treatment of intravenous extravasations: implementation of an evidence-based guideline in a pediatric population. J Spec Pediatr Nurs. 2013;18(3):253–262. doi: https://doi.org/10.1111/jspn.12035

32. Boyar V, Galiczewski C. Efficacy of Dehydrated Human Amniotic Membrane Allograft for the Treatment of Severe Extravasation Injuries in Preterm Neonates. Wounds. 2018;30(8):224–228.

33. Yan Y, Gong M, Chen J, et al. Incidence, risk factors and treatment outcomes of drug extravasation in pediatric patients in China. Turk J Pediatr. 2017;(59):162–168.

34. Myers P, Krasniak P, Bell D. Managing intravenous infiltration injuries in the neonatal intensive care unit. J Burn Care Res. 2018;39(9):S151.

35. Loth T, Eversmann W. Extravasation injuries in the upper extremity. Clin Orthop Relat Res. 1991;(272):48.

36. Scuderi N, Onesti M. Antitumor agents: extravasation, management, and surgical treatment. Ann Plast Surg. 1994;32(1):39.

37. Langstein N, Duman H, Seelig D, et al. Retrospective study of the management of chemotherapeutic extravasation injury. Ann Plast Surg. 2002;49(4):369.

38. D’Andrea F, Onesti M, Nicoletti G, et al. Surgical treatment of ulcers caused by extravasation of cytotoxic drugs. Scand J Plast Reconstr Surg Hand Surg. 2004;38(5):288–292.

39. Rose R, Felix R, Crawford-Sykes A, et al. Extravasation injuries. West Indian Med J. 2008;57(1):40.

40. Sbitany H, Koltz F, Mays C, et al. CT contrast extravasation in the upper extremity: strategies for management. Int J Surg. 2010;8(5):384.

41. Kim S, Cook K, Lee I, et al. Computed tomography contrast media extravasation: treatment algorithm and immediate treatment by squeezing with multiple slit incisions. Int Wound J. 2017;14(2):430–434.

42. Onesti M, Carella S, Fioramonti P, et al. Chemotherapy Extravasation Management: 21- Year Experience. Ann Plast Surg. 2017;79(5):450–457.

43. Millam D. Managing complications of IV therapy. Nursing (Lond). 1988;(18):34-43.

44. Flemmer L, Chan J. A pediatric protocol for management of extravasation injuries. Pediatr Nurs. 1993;18:44–47.

